# Underdetection of COVID-19 cases in France in the exit phase following lockdown

**DOI:** 10.1101/2020.08.10.20171744

**Authors:** Giulia Pullano, Laura Di Domenico, Chiara E Sabbatini, Eugenio Valdano, Clément Turbelin, Marion Debin, Caroline Guerrisi, Charly Kengne-Kuetche, Cécile Souty, Thomas Hanslik, Thierry Blanchon, Pierre-Yves Boëlle, Julie Figoni, Sophie Vaux, Christine Campèse, Sibylle Bernard-Stoecklin, Vittoria Colizza

## Abstract

A novel testing policy was implemented in May in France to systematically screen potential COVID-19 infections and suppress local outbreaks while lifting lockdown restrictions. 20,777 virologically-confirmed cases were notified in mainland France from May 13, 2020 (week 20, end of lockdown) to June 28 (week 26). Accounting for missing data, positive tests before symptom onset, and the delay from symptom onset to test, this corresponds to 14,061 [95% CI 13,972-14,156] cases with symptom onset during this period, a likely underestimation of the real number. Using age-stratified transmission models parameterized to behavioral data and calibrated to regional hospital admissions, we estimated that 103,907 [95% CI 90,216-116,377] COVID-19 symptomatic cases occurred, suggesting that 9 out of 10 cases with symptoms were not ascertained. Median detection rate increased from 7% [6-8]% to 38% [35-44]% over time, with large regional variations. Healthcare-seeking behavior in COVID-19 suspect cases remained low (31%). Model projections for the incidence of symptomatic cases (6.7 [5.8, 7.4] per 100,000 in week 26) were compatible with estimates from participatory surveillance, provided that 80% of suspect cases consulted. Encouraging awareness and same-day healthcare-seeking behavior in suspect cases is critical to improve detection. The capacity of the system remained however insufficient even at the low levels of viral circulation achieved after lockdown, and was predicted to deteriorate rapidly for increasing epidemic activity. Substantially more aggressive, targeted, and efficient testing with easier access is required to act as a pandemic-fighting tool. These elements should be considered in light of the currently observed increase of cases in France and other European countries.

---

As countries in Western Europe gradually relaxed lockdown restrictions, robust surveillance and detection systems became critical to monitor the epidemic situation and maintain activity at low levels^1^. The need is to rapidly identify and isolate cases to prevent onward transmission in the community and avoid substantial resurgence of cases. In France, the surveillance strategy implemented by authorities to exit lockdown on May 11, 2020 was multifold^2^ and based on an expanded case definition for COVID-19 suspect cases to guide clinical diagnosis^3^; recommendations to the general population to seek healthcare even in presence of mild symptoms; prescription of diagnostic tests to suspect cases by general practitioners for systematic and comprehensive testing; isolation of confirmed cases and tracing of their contacts.

The specific characteristics of COVID-19 epidemic, however, hinder the identification of cases^4^. Large proportions of asymptomatic infectious individuals^5^, and presence of mild or paucisymptomatic infections that easily go unobserved^6,7^ present serious challenges to detection and control^7–9^. This may potentially result in substantial underestimates of the real number of COVID-19 cases in the country. Here we estimated the rate of detection of COVID-19 symptomatic cases in France in May-June 2020 after lockdown, through the use of virological and participatory syndromic surveillance data coupled with mathematical transmission models calibrated to regional hospitalizations. The study focused on mainland France where the epidemic situation was comparable across regions, and excluded Corsica reporting a very limited epidemic activity and overseas territories characterized by increasing transmission^10^.

## COVID-19 surveillance

COVID-19 epidemic management in France in the post-lockdown phase involved the creation of a centralized database collecting data on virological testing (SI-DEP, Information system for testing) to provide indicators to monitor the epidemic over time^2,11^. All individuals with symptoms compatible with COVID-19 were invited to consult their general practitioner and obtain a prescription for a virological test. Contacts of confirmed cases were traced and tested. SI-DEP centralizes results of all virological tests conducted in France. 20,777 virologically-confirmed cases were notified from May 13 (week 20) to June 28 (week 26) in mainland France. These include positive individuals with or without symptoms at the time of testing, or positive individuals for which information on clinical status at the time of testing was missing. Accounting for presymptomatic individuals among those presenting with no symptoms at the time of testing and after imputation of missing data (see Methods), an estimated 16,165 [95%CI 16,101-16,261] symptomatic cases were tested in the study period (**Figure 1**). The average delay from symptom onset to testing decreased from 12.5 days in week 20 (w20) to 2.8 days in w26. Accounting for this delay (see Methods), we estimated that 14,061 [13,972-14,156] confirmed symptomatic cases had onset of symptoms in the study period, showing a decreasing trend over time (2,493 in w20, 1,647 in w26). The test positivity rate decreased in the first weeks and stabilized around 1.2% (average over w24-w26).

**Figure 1.**
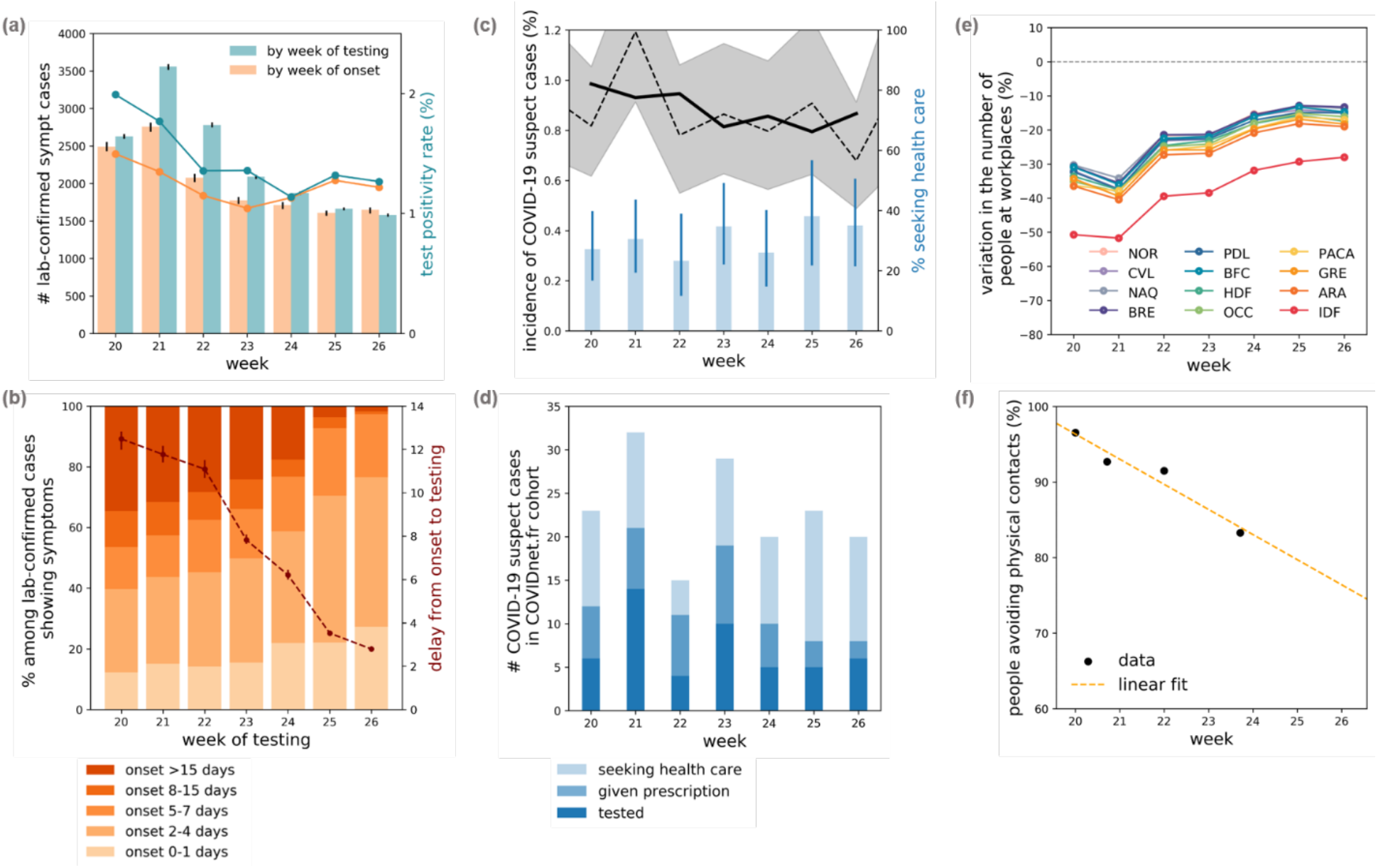
Virological surveillance, participatory syndromic surveillance, behavioral data for model parameterization. (a) Estimated number of virologically-confirmed symptomatic cases in mainland France by week of testing and week of onset, along with the test positivity rate. Estimates are based on imputation of positive individuals without symptoms at the time of testing into asymptomatic or presymptomatic; imputation of missing data on clinical status at the time of testing into asymptomatic, presymptomatic, or symptomatic; imputation of the date of onset of symptoms for presymptomatic and symptomatic cases. Uncertainties correspond to 95% CI. Week of symptom onset for symptomatic cases was estimated based on patients’ declarations (see panel b) through a monthly Gamma distribution fitted to the data with a maximum likelihood approach. Missing data about presence/absence of symptoms were imputed by region and by week, by sampling from a multinomial distribution according to the observed breakdown among cases with complete information (see Methods). Test positivity rate was computed on cases with complete information. Data for weeks 20-26 were consolidated in w30. (b) Breakdown of virologically-confirmed cases with symptoms and complete information in SI-DEP database by week of testing according to declared onset of symptoms. Estimated time from onset to testing is also shown (right y axis, median and 95% CI). (c) Incidence of COVID-19 suspect cases (estimates by week, median and 95% CI, and 3-week moving average (thick line)), along with percentage of those seeking healthcare (median and 95% CI), estimated from participatory surveillance system COVIDnet.fr. (d) Number of COVID-19 suspect cases of the participatory cohort seeking healthcare, and among them those receiving a prescription, and performing a virological test given the prescription. COVIDnet.fr estimates were adjusted on age and sex of participants. (e) Estimated change in individuals’ presence at workplace locations over time and by region based on Google location history data^14^. Region acronyms are listed in Table 1. (f) Percentage of individuals avoiding physical contacts with respect to lockdown estimated from a large-scale survey conducted by Sante publique France^15^.

A digital participatory system was additionally considered for COVID-19 syndromic surveillance in the general population^10^, including those who do not consult a doctor. Called COVIDnet.fr, it was adapted from the platform GrippeNet.fr (dedicated to influenza-like-illness surveillance since 2011^12,13^) to respond to the COVID-19 health crisis in early 2020. It is based on a set of volunteers who weekly self-declare their symptoms, along with socio-demographic information. Based on symptoms declared by an average of 7,500 participants each week, the estimated incidence of COVID-19 suspect cases decreased from about 1% to 0.8% over time (**Figure 1**), according to the expanded suspect case definition recommended by the High Council of Public Health for testing^3^ (Methods). 162 out of 524 suspect cases (31%) consulted a doctor in the study period. Among them, 89 (55%) received a prescription for a test, resulting in screening for 50 individuals (56% of those given the prescription).

**Table 1.**
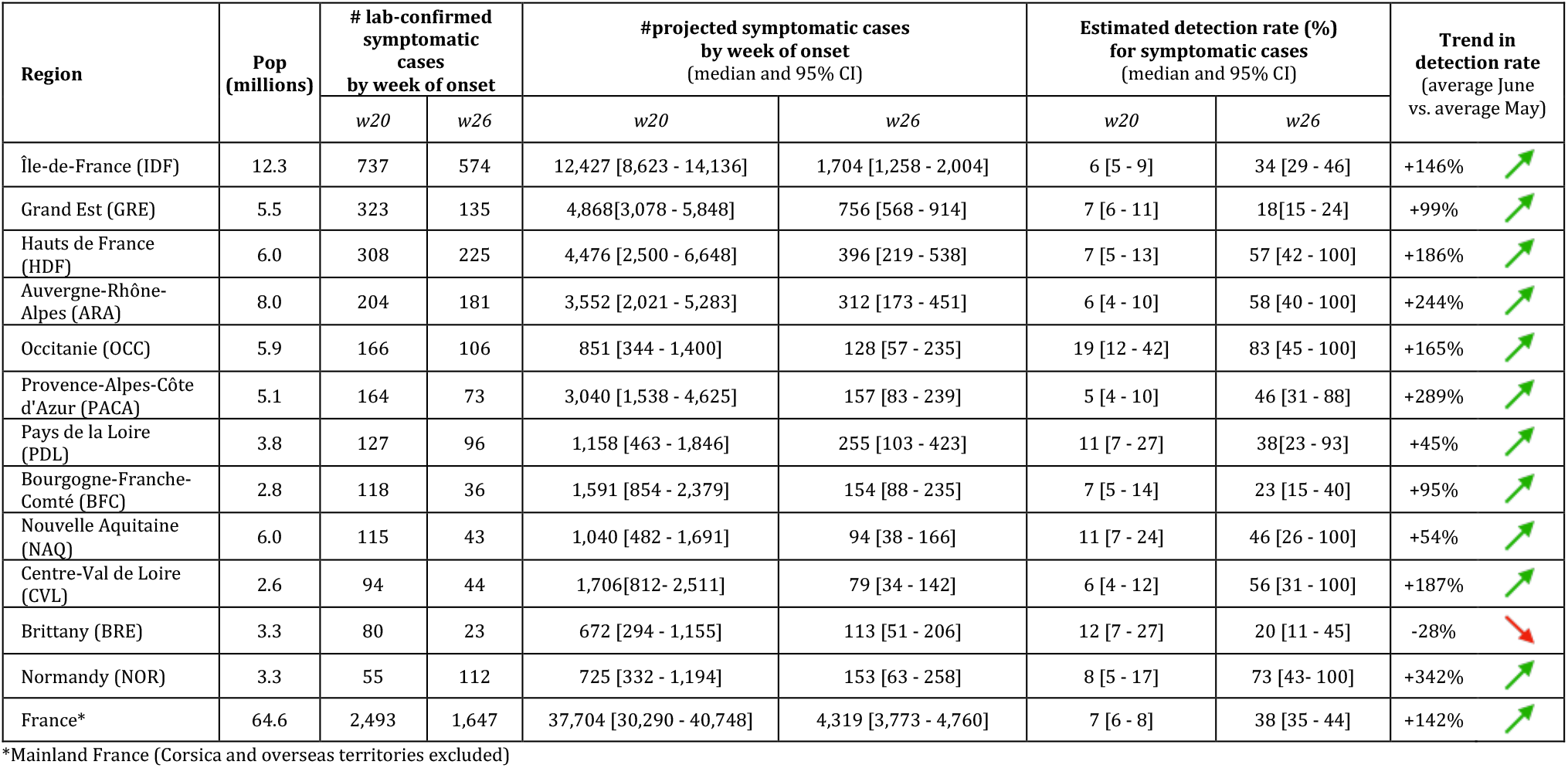
Number of virologically-confirmed symptomatic cases, number of projected symptomatic cases, estimated detection rate, estimated trend in detection rate, population per region. Regions are ranked by decreasing number of confirmed cases in w20. The trend is estimated comparing the average of the estimated detection rate in the weeks of June (w23-26) with the average in the weeks of May (w20-w22).

| Region | Pop (millions) | # lab-confirmed symptomatic cases by week of onset |  | #projected symptomatic cases by week of onset (median and 95% CI) |  | Estimated detection rate (%) for symptomatic cases (median and 95% CI) |  | Trend in detection rate (average June vs. average May) |
| --- | --- | --- | --- | --- | --- | --- | --- | --- |
|  |  | w20 | w26 | w20 | w26 | w20 | w26 |  |
| Île-de-France (IDF) | 12.3 | 737 | 574 | 12,427 [8,623 - 14,136] | 1,704 [1,258 - 2,004] | 6 [5 - 9] | 34 [29 - 46] | +146% |
| Grand Est (GRE) | 5.5 | 323 | 135 | 4,868[3,078 - 5,848] | 756 [568 - 914] | 7 [6 - 11] | 18[15 - 24] | +99% |
| Hauts de France (HDF) | 6.0 | 308 | 225 | 4,476 [2,500 - 6,648] | 396 [219 - 538] | 7 [5 - 13] | 57 [42 - 100] | +186% |
| Auvergne-Rhône-Alpes (ARA) | 8.0 | 204 | 181 | 3,552 [2,021 - 5,283] | 312 [173 - 451] | 6 [4 - 10] | 58 [40 - 100] | +244% |
| Occitanie (OCC) | 5.9 | 166 | 106 | 851 [344 - 1,400] | 128 [57 - 235] | 19 [12 - 42] | 83 [45 - 100] | +165% |
| Provence-Alpes-Côte d'Azur (PACA) | 5.1 | 164 | 73 | 3,040 [1,538 - 4,625] | 157 [83 - 239] | 5 [4 - 10] | 46 [31 - 88] | +289% |
| Pays de la Loire (PDL) | 3.8 | 127 | 96 | 1,158 [463 - 1,846] | 255 [103 - 423] | 11 [7 - 27] | 38[23 - 93] | +45% |
| Bourgogne-Franche-Comté (BFC) | 2.8 | 118 | 36 | 1,591 [854 - 2,379] | 154 [88 - 235] | 7 [5 - 14] | 23 [15 - 40] | +95% |
| Nouvelle Aquitaine (NAQ) | 6.0 | 115 | 43 | 1,040 [482 - 1,691] | 94 [38 - 166] | 11 [7 - 24] | 46 [26 - 100] | +54% |
| Centre-Val de Loire (CVL) | 2.6 | 94 | 44 | 1,706[812 - 2,511] | 79 [34 - 142] | 6 [4 - 12] | 56 [31 - 100] | +187% |
| Brittany (BRE) | 3.3 | 80 | 23 | 672 [294 - 1,155] | 113 [51 - 206] | 12 [7 - 27] | 20 [11 - 45] | -28% |
| Normandy (NOR) | 3.3 | 55 | 112 | 725 [332 - 1,194] | 153 [63 - 258] | 8 [5 - 17] | 73 [43 - 100] | +342% |
| France* | 64.6 | 2,493 | 1,647 | 37,704 [30,290 - 40,748] | 4,319 [3,773 - 4,760] | 7 [6 - 8] | 38 [35 - 44] | +142% |

\*Mainland France (Corsica and overseas territories excluded)

### Projections of COVID-19 epidemic trajectories and estimated detection rates

We used stochastic discrete age-stratified epidemic models^16,17^ based on demography, age profile^18^, and social contact data^19^ of the 12 regions of mainland France, to account for age-specific contact activity and role in COVID-19 transmission. Disease progression is specific to COVID-19^16,17^ and parameterized with current knowledge to include presymptomatic transmission^20^, asymptomatic^5^ and symptomatic infections with different degrees of severity (paucisymptomatic, with mild symptoms, with severe symptoms requiring hospitalization)^7,21–23^. The model was shown to capture the transmission dynamics of the epidemic in Île-de-France and was used to assess the impact of lockdown and exit strategies^16,17^. Full details are reported in the Methods section and in the Supplementary Material.

Intervention measures were modeled as mechanistic modifications of the contact matrices, accounting for a reduction of the number of contacts engaged in specific settings, and were informed from empirical data (see Methods). Lockdown data came from Refs.^16,17^. The exit phase was modeled considering region-specific data of attendance at school based on Ministry of Education’s data^24^, partial maintenance of telework according to estimated presence in workplaces from mobile phones location history data^14^ (**Figure 1**), reduction in adoption of physical distancing over time based on survey data^15^ (**Figure 1**), senior protection^15^, partial reopening of activities. A sensitivity analysis was performed on the reopening of activities, as data were missing for an accurate parameterization of associated contacts. Testing and isolation of detected cases were implemented by considering a 90% reduction of contacts for the number of virologically-confirmed COVID-19 cases^16,17^. Region-specific models were fitted to regional hospital admission data (**Figure 2**) through a maximum likelihood approach in the phases before lockdown, during lockdown, and in the exit phase. Further details are reported in the Methods section and Supplementary Material.

**Figure 2.**
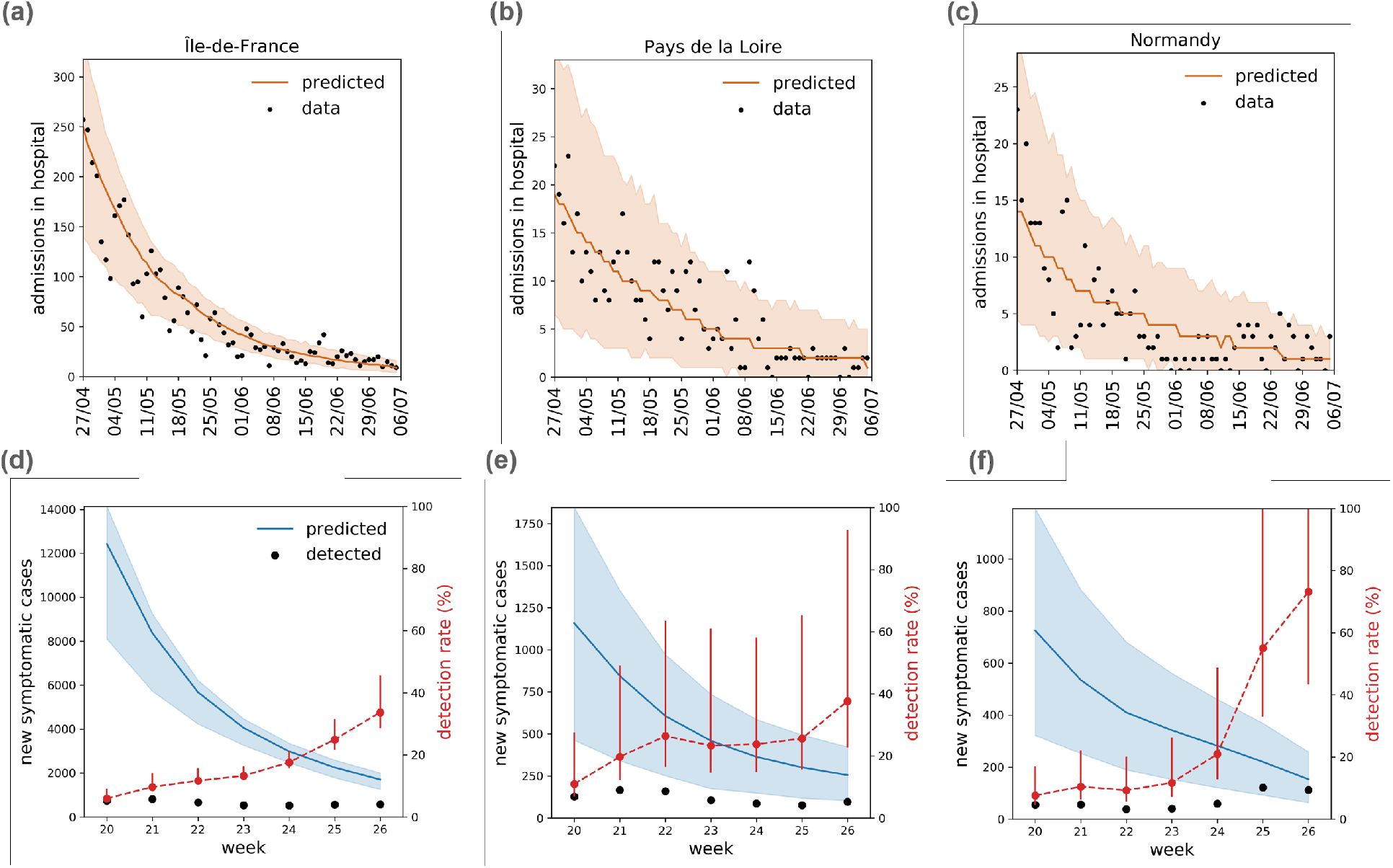
Hospital admissions and number of new symptomatic cases. (a-c) Hospital admissions over time, data (points) and simulations (median and 95% CI), for Île-de-France (a), Pays de la Loire (b), Normandie (c). Hospital admission data up to w27 (consolidated in w28) were used to infer parameter values. (d-f) Projected number of new symptomatic cases over time (median and 95% CI) and estimated number of virologically-confirmed symptomatic cases by week of onset (points), for the same regions above. The estimated detection probability of symptomatic cases (%) is also shown (red points, median and 95% CI, right y axis). Plots for the remaining regions are reported in the Supplementary Material.

Projected number of cases decreased over time in all regions, in agreement with the decreasing tendency reported in hospital admissions in the study period (**Figure 2**). Overall, 103,907 [95% CI 90,216-116,377] new infections were predicted in mainland France in weeks 20-26 (from 37,704 [30,290-40,748] in w20 to 4,319 [3,773-4,760] in w26). Île-de-France was the region with the largest predicted number of cases (12,427 [8,623-14,136] to 1,704 [1,258 -2,004] from w20 to w26), followed by Grand Est and Hauts-de-France (**Table 1, Table S3** of the Supplementary Material).

Projections were substantially higher than virologically-confirmed cases (**Figures 2** and **3**). The estimated detection rate for symptomatic infections in mainland France in the period w20-w26 was 14% [12-16]%, suggesting that slightly less than 9 out of 10 new cases with symptoms were not identified by the surveillance system. Estimated detection rate increased over time (7% [6-8]% in w20, 38% [35-44]% in w26). By the end of June, 5 regions had a median detection above 50%, and 6 regions detected a number of cases within the confidence interval of model projections (**Fig. 3b-d, Table 1**). All regions except Brittany displayed average increasing trends in the estimated detection rate in June compared to May. We did not find significant associations between the detection rate and the number of detected cases by week of onset, or the positivity rate of performed tests (Supplementary Material). However, the detection rate was negatively associated with the model-predicted incidence (Spearman correlation *r* = −0.75, *p* < 10^−15^; **Fig. 3f**). In addition, the data followed a power-law function, *π* = 66 · *i*^−0.51^, where *π* is the weekly detection rate of symptomatic cases (expressed in %) and *i* is the projected weekly incidence of symptomatic cases (expressed in cases per 100,000). This function quantifies the relation, in the study period, between the detection capacity of the test-trace-isolate system and the viral circulation in the population. It clearly shows that capacity rapidly drops as epidemic activity increases.

**Figure 3.**
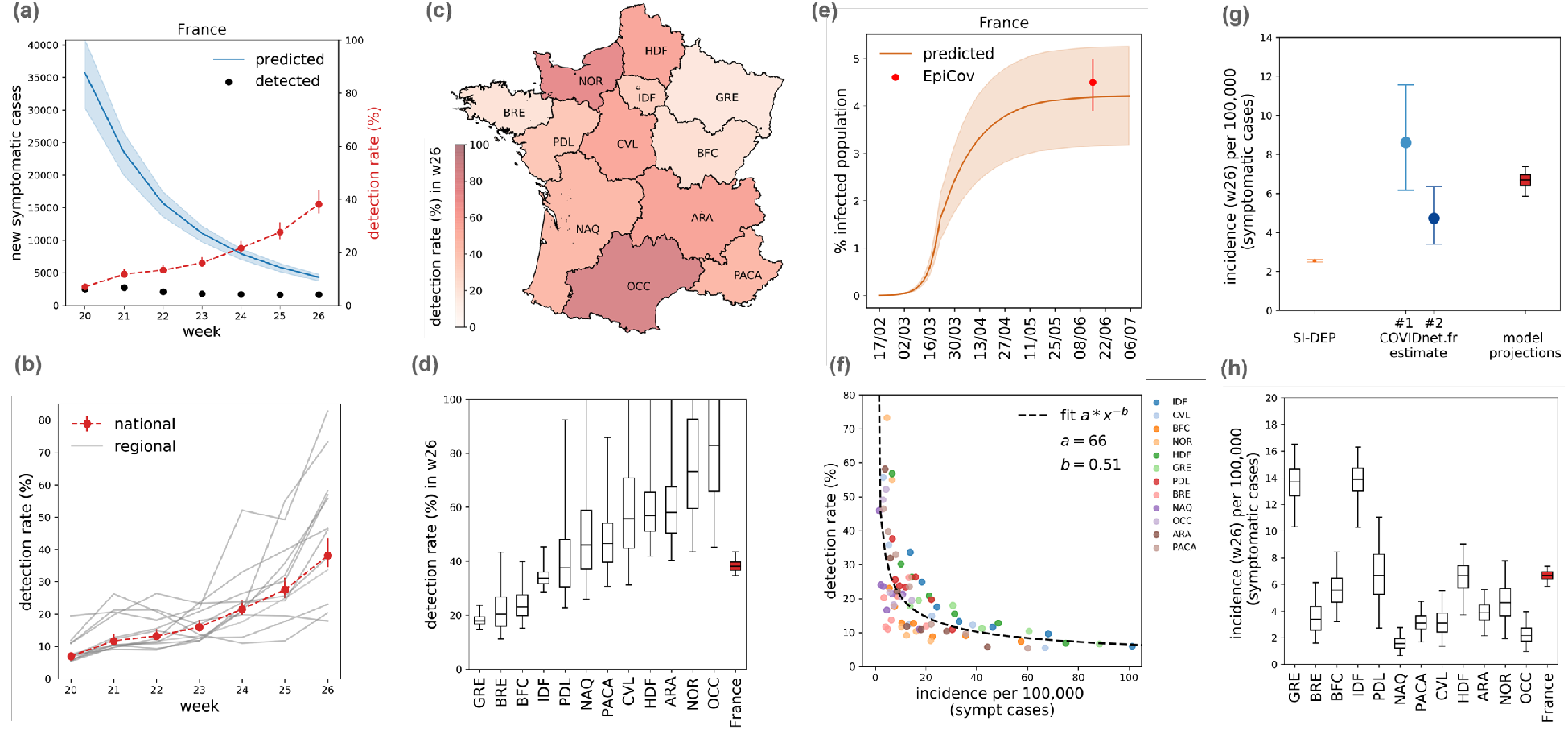
Detection rate and incidence. (a) Projected number of new symptomatic cases over time (median and 95% CI) and estimated number of virologically-confirmed symptomatic cases by week of onset (points) in mainland France. The estimated detection rate of symptomatic cases (%) is also shown (red points, median and 95% CI, right y axis). (b) Estimated detection rate of symptomatic cases (%) and 95% CI over time for mainland France (red dots), and for all regions (grey lines, where only median values are shown for the sake of visualization). (c) Map of the estimated detection rate (%) by region in w26 (June 22-28, 2020). (d) Estimated detection rate per region compared to the national estimate. Regions are ranked by increasing median detection rate. Boxplots with whiskers represent the median (line in the middle of the box), interquartile range (box limits), and 2.5^th^ and 97.5^th^ percentiles (whiskers). (e) Predicted percentage of population infected (median and 95% CI) compared with estimates from a serological study performed on a representative sample of the population in mainland France (EpiCov^27^, red dot, see Supplementary Material for more details). (f) Estimated detection rate of symptomatic cases (%) by region and by week vs. projected incidence by region and by week. The curve indicates the result of a least-square fit to the data with a power-law function, *π* = 66 · *i*^−1.01^, where *π* is the detection rate (expressed in %) and *i* is the weekly incidence (expressed in number of cases per 100,000). Errors in the parameters are reported in the Supplementary Material. (g) Estimated incidence of symptomatic cases and 95% CI in mainland France in w26 from different sources: virological surveillance data (SI-DEP), participatory surveillance data (COVIDnet.fr, with two estimates), model projections. (h) Projected incidence per region compared to the national estimate. Regions are ranked as in panel d. Boxplots with whiskers are defined as in panel d.

Validation of the model was performed in two ways. First, we compared our model projections of the percentage of infected population with the results of three independent seroprevalence studies performed after the first wave in France^25–27^. Modeling results are in agreement with serological estimates at national and regional level and at different points in time (**Fig. 3e** for the comparison with the serological study performed on a representative sample of the population, at the national level; results by region and with the other two serological studies are provided in the Supplementary Material). Second, we compared the projected incidence of COVID-19 symptomatic cases in w26 (6.69 [5.84-7.37] per 100,000) with the value obtained from virologically-confirmed cases (2.55 [2.48-2.61] per 100,000) and two estimates based on COVIDnet.fr data (**Fig. 3g**). The first estimate applies the measured test positivity rate to the incidence of self-reported COVID-19 suspect cases (estimate #1, yielding 8.6 [95% CI 6.2-11.5] per 100,000); the second additionally assumes that only 55% would be confirmed as suspect case by a physician and prescribed a test (according to COVIDnet.fr data on consulting participants, estimate #2, yielding 4.7 [3.4-6.3] per 100,000). Our projections are in line with plausible estimates from COVIDnet.fr, and suggest that on average at least 80% of suspect cases should be tested to recover the predicted incidence.

Sensitivity analysis showed that findings were robust against elements of the contact matrices that could not be informed by empirical data (Supplementary Material). Also, a model selection analysis showed that changes in the contact patterns over time due to restrictions and activities of individuals of different age classes during the exit phase (e.g. for partial attendance at school, telework) are needed to accurately capture the transmission dynamics (Supplementary Material).

The detection rate of asymptomatic cases was computed assuming a delay from infection to test based on the infection-to-onset distribution of the compartmental model and onset-to-test delay of symptomatic individuals (see Methods), as no additional information on the cases was available to inform the date of infection. We found lower detection rates for asymptomatic individuals, ranging from 2.8% [2.5-3.3]% in w20 to 14.7% [13.2-16.8]% in w26 for mainland France (see also Supplementary Material).

## Discussion

Despite a test positivity rate in mainland France well below WHO recommendations (5%)^28^, a substantial proportion of symptomatic cases (9 out of 10) remained undetected in the first 7 weeks following the end of lockdown. Around 90,000 symptomatic infections were not ascertained by the surveillance system from May 11 to June 28, 2020, according to our estimates.

Surveillance improved substantially over time. Detection rate was estimated to be 7% [6-8]% at the national level in mid-May, in line with estimates for the same period from a seroprevalence study in Geneva, Switzerland^29^. By the end of June, it increased to 38% [35-44]%, leaving about 6 cases with symptoms out of 10 undetected. Six regions (Hauts de France, Auvergne-Rhône-Alpes, Occitanie, Nouvelle Aquitaine, Centre-Val de Loire, Normandy) reported cases compatible with model projections. Improvement could have resulted from the combination of different aspects. On one side, the new surveillance framework may have progressively strengthened with increasing resources and capacity over time, as signaled for example by a more rapid detection of cases (78% reduction of the delay from symptom onset to testing from May to June). On the other side, the system certainly benefited from a substantial and concurrent decrease of the epidemic activity in all regions.

Despite this positive trend, our findings highlight structural limitations and a critical need for improvement. Some regions remained with limited diagnostic exhaustiveness by the end of June. This is particularly concerning in those regions predicted to have a large number of weekly infections, such as Île-de-France where approximately only 1 out of 3 cases with symptoms were detected by the end of June, and Grand Est (2 out of 10). Novel recommendations since the end of lockdown require that all patients with symptoms suggestive of COVID-19 (as well as contacts of a confirmed case) be screened for SARS-CoV-2^2^. Almost all cases (92% since May 25) clinically diagnosed by sentinel general practitioners as possible COVID-19 cases were prescribed a test^10^. However, only 31% of individuals with COVID-19-like symptoms consulted a doctor in the study period according to participatory surveillance data. Overall, these figures suggest that a large number of symptomatic COVID-19 cases were not screened because they did not seek medical care despite recommendations. A similar evidence emerged from a large-scale serological study in Spain where only between 16% and 20% of symptomatic participants with antibodies against SARS-CoV-2 reported a previous virological screening^30^. By combining estimates from virological and participatory surveillance, we extrapolated an incidence of symptomatic cases from crowdsourced data that is compatible with model projections, under the hypothesis that the large majority of suspect cases would get tested. This finding further supports testing for all COVID-19 suspect cases, and suggests that the requirement of the test prescription can be lifted. Large-scale communication campaigns should reinforce recommendations to raise awareness in the population and strongly encourage healthcare-seeking behavior especially in patients with mild symptoms. At the same time, investigations to identify reasons for not consulting could be quickly performed through the participatory surveillance system.

Red tape might have contributed to low testing rates. Prescription for a test was deemed compulsory in the new testing policy to prevent misuse of diagnostic resources^2^, however the path involving consultation, prescription, and lab appointment may have discouraged mildly affected individuals not requiring medical assistance. To facilitate access, testing should not require a prescription, as then established by authorities at the end of July^31^. In addition, some local initiatives emerged over summer that increased the number of drive-through testing facilities, mailed test vouchers to promote massive screening in certain regions (e.g. in Île-de-France^32^), offered temporary mobile testing facilities (buses, pavilions) to increase proximity with the population^33^. These initiatives are particularly relevant for counteracting socio-economic inequalities in access to information and care in populations vulnerable to COVID-19^34^. However, such strategies should not hinder a testing protocol targeting suspect index cases, and subsequent follow-up through contact tracing investigations. Our results indeed show that high testing efforts, measured by low test positivity rates, are not necessarily associated to high rates of detection. This was also observed in the UK during the first wave when detection remained low despite the large number of tests performed (and low positivity rate)^35^. Without a strong case-based surveillance, the risk is to disperse resources towards random individuals without symptoms who are unlikely to be positive. This might saturate the test-trace-isolate machinery, without achieving the low level of viral circulation required to safeguard the hospital system. The large demand for testing observed over summer in certain regions, mainly as a result of imminent travels and protocols imposed by certain countries and air companies, as well as untargeted recommendations, reportedly caused long waiting lists and overwhelmed testing sites^36^. Delays from onset to screening were still longer than 48h at the end of June, and delays in providing results increased over time, from 1-2 days to about 1 week or longer^37^. Given pre-symptomatic transmission, notification to contacts should be almost immediate to allow the effective interruption of transmission chains^20^. For testing to be an actionable tool for surveillance and, most importantly, for control of COVID-19 transmission, delays should be suppressed and screening rates radically increased and better targeted. Over May-June the average weekly number of tests was 250,000, remaining well below the objective originally fixed by authorities (700,000). In these conditions, we found that the capacity of detection of the test-trace-isolate system scaled as the inverse of the square root of the incidence, rapidly deteriorating already at low incidence levels. The system was predicted to be able to detect more than 2 cases out of 3 (rate>66%) only if the incidence was lower than 1 symptomatic case per 100,000, a figure 50 times smaller than what we projected at the exit from lockdown at the national level. This suggests that, even right after lockdown, the system was unable to perform the comprehensive surveillance of suspect cases recommended by WHO^28^ while phasing out the restrictions. More aggressive testing should be performed at low activity level to keep the epidemic under control. Failing in doing so leads to a rapid and uncontrolled increase of cases with large-scale transmission in the community^9^, as currently reported in European countries^38^. Such risk is even stronger if the relaxation in preventive behaviors persists due to adhesion fatigue^15^, as reported during summer.

The number of tests increased over time after the study period, together with changes in the testing policy (test prescription was not needed anymore). Such improvement, however, was a reactive response to the first resurgence of cases in certain regions in France in July, and thus likely counterbalanced by a larger incidence of cases. Future studies should determine if the relationship between detection and incidence that we found is robust over time, as that relationship is key to estimate the needed capacity in terms of number of tests per week, to fine-tune the testing strategy, as well as the length and intensity of the restrictive measures put in place to reduce the viral circulation to manageable levels.

Aggressive and efficient testing will become increasingly more important in the fall and winter months, as other respiratory viruses, such as influenza, RSV, rhinoviruses, will start to circulate. Reviewing the testing strategy while at low COVID-19 epidemic activity, is an important opportunity to strengthen French response system for the next season.

Models were region-based and did not consider a possible coupling between regional epidemics caused by mobility. This choice was supported by stringent movement restrictions during lockdown^39^, and by the limited mobility increase in May-June^40^, before important inter-regional displacements took place at the start of summer holidays in July. Foreign importations were neglected^8,41^ as France reopened its borders with EU Member States on June 15, and the Schengen area remained closed till July. COVIDnet.fr cohort is not representative of the general population^13^, however the agreement found with sentinel incidence trends for influenza-like-illness suggests that these limitations have little effect once results are adjusted for lack of representativeness^12^. Underdetection may also proceed from the imperfect characteristics of RT-PCR (reverse transcription-polymerase chain reaction) tests used to identify infected cases^42^. Some cases tested for SARS-CoV-2 could have been falsely negative, e.g. because tested too early after the infection. This would affect the analysis presented in the manuscript and would be in line with our conclusion that a large part of cases may have been undetected. A previous work assessed the rate of underdection of symptomatic cases in 210 countries^35^, but it mainly focused on the early global dynamics. Our model gives up geographical extension for higher data quality in a specific country, thus providing a novel synthesis of data sources characterizing human behavior over time and space with virological and participatory surveillance data. Our model clearly identifies in the targeted testing of symptomatic cases the weak link of epidemic response, which requires prompt improvement. Our projections were validated with serological estimates after the first wave and participatory surveillance estimates in the exit phase after lockdown. Model selection demonstrated the importance of data sources describing changes of human behavior. The study was not extended to the summer months, because of (i) the challenge of mechanistically parameterizing the contact matrices during summer holidays, (ii) the increase of movement fluxes across regions weakening our assumption of region-specific models, and (ii) the interruption of COVIDnet.fr surveillance during the summer break preventing the identification of the key factors behind case underascertainment.

Our findings identify critical needs of improvement to increase case ascertainment in France and the performance of the response system to monitor and control COVID-19 epidemic. Substantially more aggressive and efficient testing targeting COVID-19 suspect cases needs to be achieved to act as a pandemic-fighting tool. Associated logistical needs should not be underestimated. These elements should be considered in light of the increase of cases currently observed in France and in other countries in Europe with similar response systems.

## Methods

### Virological surveillance data

The centralized database SI-DEP for virological surveillance^11^ collects all tests performed in France for any reason. In the period under study, guidelines recommended to consult a general practitioner at the first sign of COVID-19-like symptoms and obtain a prescription for a virological test (prescription was compulsory to access the test). In addition, routine testing was performed for patients admitted to the hospital with any diagnosis, healthcare personnel, and individuals at other facilities (e.g. in some senior homes or long-term healthcare facilities). Data report detailed information on individuals tested in France, including (i) date of test, (ii) result of test (positive or negative), (iii) location (region), (iv) absence or presence of symptoms at the time of testing, (v) self-declared delay between onset to test in presence of symptoms. The delay is provided with the following breakdown: onset date occurring 0-1 day before date of test, 2-4 days before, 5-7 days before, 8-15 days before, or >15 days before. For some tests, information on (iv) and (v) is missing. The SI-DEP database provided complete information for 23,210 (66%) out of 35,264 laboratory-confirmed COVID-19 cases tested between week 20 (May 11-May 17) and week 30 (July 19-July 26), with an increasing trend of complete information over time (see Supplementary Material). The study referred to the period from w20 to w26. Data up to w30 were used to consolidate the data in the study period accounting for the delays.

### Imputation of asymptomatic vs. presymptomatic cases, of onset date, and of missing information

Individuals who tested positive on a given date were recorded in the SI-DEP database as: cases with symptoms at the time of testing, with a self-declared delay from onset of symptoms; cases without symptoms at the time of testing; or cases with no information on presence/absence of symptoms at the time of testing. These three subsets of cases were analysed to account for presence of presymptomatic individuals among those with no symptoms at time of testing, imputation of missing data, estimation of dates of infection or symptom onset.

- For laboratory-confirmed COVID-19 cases with symptoms at the time of testing, we estimated their date of onset using the information on the date of test and the time-interval of onset-to-test delay self-declared by the patients (**Fig. 1b**). We fitted a Gamma distribution to onset-to-test delay data with a maximum likelihood approach, using three different periods of time (May, June, July), to account for changes in the distribution of self-declared delays over time (i.e. longer delays at the beginning of the study period, shorter delays at its end). We obtained average delays of 12.9 days in May, 5.1 days in June, 2.7 days in July (Supplementary Material). July data was used to consolidate data corresponding to infections with onset in June and tested with delay. Given a confirmed case with symptoms testing on a specific date, we assigned the onset date by sampling the onset-to-testing delay from the fitted distribution for that period, conditional to the fact that the delay lies in the corresponding time-interval declared by the patient. The imputation procedure was carried out 100 times. Results were aggregated by week of onset.
- For laboratory-confirmed COVID-19 cases with no symptoms at the time of testing, we assumed that on average 40% of them were asymptomatic^5^ (see transmission model subsection), whereas the remaining 60% were presymptomatic who tested early thanks to contact tracing. Imputation was done by sampling from a binomial distribution and repeated 100 times. Data on contact tracing could not be used to inform data on infection or symptom onset, because of national regulatory framework on privacy. Given the low sensitivity of PCR tests in the early phase of the incubation period, we considered that imputed presymptomatic cases belonged to the prodromic phase, and estimated their onset date based on the structure of our compartmental model. For imputed asymptomatic, we assumed the same delay from infection to testing as in cases with symptoms (see Supplementary Material). Imputation of the dates were repeated 100 times.
- For laboratory-confirmed COVID-19 cases with no information on symptoms at the time of testing, missing data were imputed by sampling from a multinomial distribution with probabilities equal to the rate of occurrence of the outcomes (asymptomatic, presymptomatic, or symptomatic with 5 possible time-intervals for the onset-to-test delay) reported for cases with complete information, and assuming the imputation of cases without symptoms into asymptomatic and presymptomatic, as described above. Imputation was performed by region and by week and repeated 100 times. Presymptomatic and symptomatic individuals were aggregated together by onset date to estimate the rate of detection of symptomatic cases.

### Participatory surveillance data and analysis

COVIDnet.fr is a participatory online system for the surveillance of COVID-19, available at www.covidnet.fr. It was adapted from GrippeNet.fr to respond to the COVID-19 health crisis in March 2020. GrippeNet.fr is a participatory system for the surveillance of influenza-like-illness available in France since 2011 through a collaboration between Inserm, Sorbonne Université and Sante publique France, supplementing sentinel surveillance^12,13,43^. The system is based on a dedicated website to conduct syndromic surveillance through self-reported symptoms volunteered by participants resident in France. Data are collected on a weekly basis; participants also provide detailed profile information at enrollment. In addition to tracking influenza-like-illness incidence^12,43^, GrippeNet.fr was used to estimate vaccine coverage in specific subgroups^44^ individual perceptions toward vaccination^45^ and health-seeking behavior^46^. It was also used to assess behaviors and perceptions related to other diseases beyond influenza^47^.

Participants are on average older and include a larger proportion of women compared to the general population^13,48^. Participating population is however representative in terms of health indicators such as diabetes and asthma conditions. Despite these discrepancies, trends of estimated influenza-like-illness incidence from GrippeNet.fr reports compared well with those of the national sentinel system^12,43^. All analyses were adjusted by age and sex of participants.

To monitor COVID-19 suspect cases in the general population, we used the expanded case definition recommended by the High Council of Public Health for systematic testing and described in their 20 April 2020 notice^3^:

- (Sudden onset of symptoms OR sudden onset of fever) AND (fever OR chills) AND (cough OR shortness of breath OR (chest pain AND age > 5 years old))
- OR (Sudden onset of symptoms OR (sudden onset of fever AND fever)) AND
  - (age > 5 years old AND (feeling tired or exhausted OR muscle/joint pain OR headache OR (loss of smell WITHOUT runny/blocked nose) OR loss of taste)
  - OR ((Age ≥ 80 years old OR Age < 18 years old) AND diarrhea)
  - OR (Age < 3 months old AND (fever WITHOUT other symptoms))).

Figure 3 reports two independent estimates obtained from COVIDnet.fr cohort data for the incidence of symptomatic cases in w26. They are computed as follows:

- *Estimate #1* = (COVIDnet.fr estimated incidence in w26) * (test positivity rate from SI-DEP in w26)
- *Estimate #2* = (COVIDnet.fr estimated incidence in w26) * (estimated proportion screened and confirmed as COVID-19 suspect case by a physician, and prescribed a test; estimates from COVIDnet.fr) * (test positivity rate from SI-DEP in w26)

The two estimates were used to validate model projections and identify the specific surveillance mechanisms needing improvement.

### Ethics statement

GrippeNet.fr/COVIDnet.fr was reviewed and approved by the French Advisory Committee for research on information treatment in the health sector (i.e. CCTIRS, authorization 11.565), and by the French National Commission on Informatics and Liberty (i.e. CNIL, authorization DR-2012–024) – the authorities ruling on all matters related to ethics, data, and privacy in the country.

### Transmission models summary

We used a stochastic discrete age-stratified transmission model for each region based on demographic, contact^19^, and age profile data of French regions^18^. Models were region-specific to account for the geographically heterogeneous epidemic situation in the country and given the mobility restrictions limiting inter-regional movement fluxes. Four age classes were considered: [0-11), [11-19), [19-65), and 65+ years old, referred to as children, adolescents, adults, seniors. Transmission dynamics follows a compartmental scheme specific for COVID-19, where individuals were divided into susceptible, exposed, infectious, and hospitalized (Supplementary Material). We did not consider further progression from hospitalization (e.g. admission to ICU, recovery, death^16^) as it was not needed for the objective of the study. The infectious phase is divided into two steps: a prodromic phase (*I*_*p*_) and a phase where individuals may remain either asymptomatic (*I*_*a*_, with probability *p*_*a*_ =40%^5^) or develop symptoms. In the latter case, we distinguished between different degrees of severity of symptoms, ranging from paucisymptomatic (*I*_*ps*_), to infectious individuals with mild (*I*_*ms*_) or severe (*I*_*ss*_) symptoms. Prodromic, asymptomatic and paucisymptomatic individuals have a reduced transmissibility *r*_*β*_ = 0.55, as estimated in Ref.^6^, and in agreement with evidence from the field^49,50^. A reduced susceptibility was considered for children and adolescents, along with a reduced relative transmissibility of children, following available evidence from household studies, contact tracing investigations, and modeling works^51–56^. A sensitivity analysis was performed on relative susceptibility and transmissibility of children, and on the proportion of asymptomatic infections (Supplementary Material). Full details are reported in the Supplementary Material.

### Contact matrices

Age-stratified transmission uses a social contact matrix that reports the average contact rates between different age classes in France^19^. This refers to the baseline condition, i.e. pre-lockdown. The contact matrix includes the following layers: contacts at home, school, workplace, transport, leisure activities, and other activities, and discriminates between physical and non-physical contacts. To account for the change of contact patterns over time, contact matrices are mechanistically parameterized, by region and over time, with different data sources informing on the number of students going to school^24^, the number of workers going to the workplace^14^, the compliance to preventive measures^15^, with a higher compliance registered in senior individuals^15^. Information on the progressive reopening of activities indicates that leisure and other activities were only partially open in the study period. Data, however, are not fine-grained enough to parameterize our model, so we assume a 50% opening of these activities and explore variations in the sensitivity analysis. Contacts in these settings are proportionally reduced according to attendance data; physical contacts are proportionally reduced according to adoption data. The parameterization of matrices is fully explained in the Supplementary Material.

### Inference framework

The parameters of the transmission models to be estimated are specific to each pandemic phase:

- Prior to lockdown, {*β, t*_0_} where *β* is the transmission rate per contact and *t*_0_ the date of the start of the simulation, seeded with 10 infectious individuals.
- During lockdown, {*α*_*LD*_, *t*_*LD*_} where *α*_*LD*_ is the scaling factor of the transmission rate per contact and *t*_*LD*_ the date when lockdown effects on hospitalization data became visible.
- After lockdown, {*α*_*exit*_ *π*_*a*_(*w*), *π*_*s*_(*w*)} where *α*_*exit*_ is the scaling factor of the transmission rate per contact, *π*_*a*_(*w*), *π*_*s*_(*w*) are the proportion of asymptomatic and symptomatic cases tested in week *w* of the exit phase, respectively. Detected cases in the simulations had their contacts reduced by 90% to mimic isolation, as done in previous work^16,17^.

We used simulations of the stochastic model to predict values for all quantities of interest (500 simulations each time). We fitted the model to the daily count of hospitalisations *H*_obs_(*d*) on day *d* throughout the period and the number of persons testing positive by week of onset, split according to disease status (symptomatic or asymptomatic), denoted *Test*_*s,obs*_(*w*) and *Test*_*a,obs*_(*w*) in week *w* of the exit phase. We used hospital admission data up to week 27 (June 29-July 5) to account for the average delay from infection to hospitalization. Data in week 27 were consolidated by waiting for one additional week to account for updates and missing data (week 28, July 6-12, 2020).

We assumed a Poisson distribution for hospitalisations and a binomial distribution for the number of people getting the test, therefore the likelihood function is

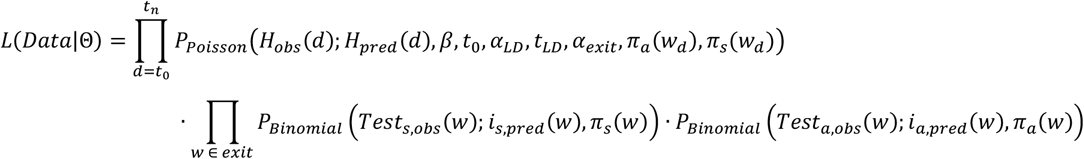

where Θ = {*β, t*_0_, *α*_*LD*_, *t*_*LD*_, *α*_*exit*_, {*π*_*a*_(*w*)}, {*π*_*s*_(*w*)}A indicates the set of parameters to be estimated, *H*_*pred*_(*d*) is the model-predicted number of hospital admissions on day *d, i*_*s, pred*_ (*w*) and *i*_*a, pred*_ (*w*) are the model-predicted weekly incidences of symptomatic and asymptomatic cases in week *w* of the exit phase, *P*_*poisson*_ is the probability mass function of a Poisson distribution, *P*_*Binomial*_ for a binomial distribution, [*t*_0_, *t*_*n*_] is the time window considered for the fit, and *w* is the week in the exit phase (w20-w26).

We reduced the required computations by making the optimization in 2 steps, first maximizing the likelihood function in the pre-lockdown and lockdown phase to estimate the first four parameters, and then maximizing the likelihood in the exit phase by fixing the first four parameters describing the epidemic trajectory prior to the exit phase to their MLEs. This second step was further simplified through an iterative procedure, and we showed through simulations that the simplified optimization procedure is consistent and well defined. The parameter space was explored using NOMAD software^57^. Fisher’s information matrix was estimated at the MLE value to obtain the corresponding confidence intervals. Simulations were then parameterized with 500 parameter sets obtained from the joint distribution of transmission parameters at MLE (one stochastic simulation for each parameter set).

Full details on the different steps and the tests performed are reported in the Supplementary Material.

### Model selection analysis

To assess the role of the mechanistic modification of the contact matrix informed by the different data sources in the exit phase, we compared our model with a simplified version assuming that contact patterns in the exit phase do not change from pre-epidemic conditions, and that all changes in the epidemic trajectory are explained exclusively by the transmissibility per contact. This is equivalent to normalize the contact matrix to its largest eigenvalue and estimate the reproductive ratio over time. We compared the two models with the Akaike information criterion.

## Data Availability

All data are available at the references cited.

## Acknowledgments

This study was partially supported by ANR project DATAREDUX (ANR-19-CE46-0008-03) and EVALCOVID-19 (ANR-20-706 COVI-0007); EU H2020 grants MOOD (H2020-874850) and RECOVER (H2020-101003589); REACTing COVID-19 modeling and surveillance grants. We thank Pascal Crepey, Camille Pelat, Edouard Chatignoux, Juliette Paireau, Daniel Levy-Bruhl for useful discussions. We also thank all participants of COVIDnet.fr system.

